# PrEP in the key population community: a qualitative study of perspectives on pre-exposure prophylaxis by men who have sex with men and female sex workers in Kigali, Rwanda

**DOI:** 10.1101/2024.08.20.24311545

**Authors:** Jonathan Ross, Josephine Gasana, Natalia Zotova, Giovanni Ndabakuranye, Fabiola Mabano, Charles Ingabire, Adebola Adedimeji, Gad Murenzi, Viraj V. Patel

## Abstract

**Background:** Men who have sex with men (MSM) and female sex workers (FSW) are increasingly and disproportionately impacted by HIV in sub-Saharan Africa, yet current PrEP care models in this region are not optimized for these communities. Limited data exist describing experiences and preferences of MSM and FSW with respect to accessing and using PrEP.

**Methods:** We conducted qualitative, semi-structured interviews with MSM and FSW recruited from three health centers and seven community organizations in Kigali, Rwanda. Data were analyzed using a mixed deductive and inductive approach to describe key themes related to initiating and adhering to PrEP.

**Results:** Participants included 18 MSM and 14 FSW; 12 were using PrEP at the time of interview, 9 had previously used PrEP, and 11 had never used it. Participants highlighted the central role of their social networks as key sources of information about and support for PrEP use, and described a strong motivation to use PrEP as a way to protect both themselves and their communities from HIV. While stigma and discrimination were pervasive, these were experienced differently by MSM and FSW. Participants suggested community access points that allowed more discreet and less frequent contact with health care workers as important and desired strategies to improve engagement.

**Conclusions:** These findings suggest that leveraging community resources for disseminating information about HIV prevention and delivering PrEP could contribute to successful implementation of PrEP for MSM and FSW in Rwanda and other settings in SSA.

## BACKGROUND

Nearly three-quarters of the 34 million people living with HIV reside in sub-Saharan Africa (SSA).^1^ Men who have sex with men (MSM) and female sex workers (FSWs) are among the key populations (KPs) who are at the highest risk of HIV, and are increasingly and disproportionately represented among new HIV infections in SSA and globally.^2,3,4^

Pre-exposure prophylaxis (PrEP) is highly effective in preventing HIV.^5–8^ In its 2016 guidelines, the World Health Organization recommended PrEP for all individuals at substantial risk of HIV.^9^ Currently, more than half of global PrEP users reside in SSA, where over 1 million individuals received a prescription in 2022.^10^ While most national PrEP programs in SSA prioritize KPs,^11^ current care models are not optimized for them. Thus, despite high willingness to use PrEP among MSM and FSW,^12–16^ engagement remains low.^17–19^ Evidence to date suggests that individual (e.g., health literacy, physical and sexual violence, stigma), institutional (e.g., discrimination in health settings), and structural factors (e.g., poverty, anti-homosexuality legislation) impact the opportunity cost around accessing health services and contribute to limited PrEP use.^20–24^

In February 2019, the government of Rwanda began implementing PrEP for KPs as part a comprehensive HIV prevention package. Initial efforts in Rwanda focused on FSW, MSM and serodiscordant couples; more recently efforts have been made to expand PrEP to adolescent girls and young women, index sex partners and individuals in the general population with considerable risk of HIV acquisition. Nonetheless, few data exist on experiences and practices of FSW and MSM with respect to accessing and using PrEP. To explore these relationships, we conducted a qualitative study to understand barriers to, facilitators of and preferences for PrEP use among Rwandan MSM and FSW.

## METHODS

### Study setting and population

Rwanda is a landlocked country situated in central Africa with a total population of 13 million; the largest and capital city is Kigali, with an approximate population of 1.7 million.^25^ A majority of FSW and MSM in Rwanda live in Kigali, where the HIV prevalence is higher (4.3%) than the national prevalence of 3%.^26^ HIV prevalence among KPs in Rwanda is markedly higher, estimated at 46% among FSW in Kigali and 7% among MSM.^27^

PrEP implementation in Rwanda has been rolled out in stages, with a focus on key populations as high-risk groups, particularly FSW and MSM. By the end of June 2023, the number of female sex workers and men who have sex with men and receiving PrEP medication had gradually increased from 10,078 in July 2022 to 10,789 in June 2023.^28^ Early studies have demonstrated high PrEP persistence among both MSM and FSW.^29^ Entry points into PrEP include self-referral, HIV testing sites, family planning services, STI treatment settings and referral from community-based organizations. PrEP care occurs at primary health centers, primarily within HIV clinics.

For the current study, we recruited participants from three health centers in Kigali (Busanza, Remera and Gikondo), as well as from seven community KP associations of MSM and FSW that operate in Kigali. The participating health centers are considered “friendly” and less stigmatizing by many MSM in Kigali, and have fairly robust PrEP programs.

### Participant recruitment

To recruit participants for the study, providers at the three health centers were asked to identify potential research participants, provide brief information about the study to them, and refer individuals who expressed interest in participation to study staff for eligibility screening. Similar recruitment efforts were made by staff and peers at the KP associations. We included participants who: 1) were HIV-negative by self-report; 2) self-identified as MSM or FSW; 3) were at least 18 years of age. Individuals who were unable to communicate in Kinyarwanda or unable to provide informed consent were excluded. We purposefully recruited individuals who were using PrEP at the time of the interview, had previously used PrEP but were not using it at the time of the interview, and individuals who had never used PrEP, such that the sample was approximately evenly divided between these categories. Participants received a stipend of 10,000 RWF (approximately $10) for their time.

### Data Collection

We developed a semi-structured interview guide informed by the socio-ecological model (Supplementary File). The interview guide explored individual and structural barriers to and facilitators of PrEP engagement, as well as preferences for PrEP care delivery. After obtaining written informed consent, interviews lasting approximately 60-90 minutes were conducted in Kinyarwanda language (the language most commonly spoken in Rwanda) by four research staff (CI, JG, GN, FM) trained in qualitative data collection and analysis. Interviewers had no prior relationship with participants.

All interviews were conducted in a private room with the presence of study staff only, to ensure recording quality and participant privacy. Each interview was conducted by a team of two interviewers, where one led the interview and the other took field notes. Interview quality was monitored by CI, observing early interviews and providing feedback with the data collection team and JR (principal investigator) through weekly conference calls. Transcripts were reviewed and compared to field notes to ensure that they reflected all content that arose during interviews. Interview guides were iteratively refined to clarify and further explore emerging themes relevant to implementation of PrEP uptake and retention in Rwanda.

### Data Analysis

Audio recordings were transcribed and translated into English transcripts, which were then analyzed using both inductive and deductive thematic analysis approach to describe key barriers, facilitators and preferences. Our approach was deductive in that the investigative team developed the initial coding scheme using the socioecological model to categorize common themes, and was inductive in that we iteratively refined the codebook based on emergent themes from reading the first six transcripts. Discrepancies were discussed and resolved by consensus. Using DeDoose software (20), the final coding scheme was independently applied to all 32 interviews by 4 coders (CI, JG, GN, FM), with each interview being coded by at least 2 investigators. The coding team regularly reviewed progress and discussed issues that arose, resolving them by consensus. After all interviews were coded, excerpts were reviewed, examining themes within each code as well as between codes and using the constant comparative method to identify, refine and consolidate emergent themes. Specifically, the analytic team examined “repeating ideas” within each code to identify emergent themes. Emergent themes were entered into a matrix, looking specifically at barriers to, facilitators of and preferences for PrEP use at different levels of the SEM. Throughout the analysis phase, the team regularly met to discuss and achieve consensus on emerging themes.

### Ethical considerations

The Rwanda National Ethics Committee (RNEC approval number 700/RNEC/2021) and the Institutional Review Board of the Albert Einstein College of Medicine (2020-12619) approved the study, which was conducted according to the principles expressed in the Declaration of Helsinki and is reported in accordance with Consolidated Criteria for Reporting Qualitative Research (COREQ) guidelines. Written informed consent forms were also obtained from all participants prior to study enrollment. The informed consent forms were kept in a locked office, transcripts were de-identified, and audio-recordings were destroyed after transcription.

## RESULTS

From November–December 2022, we interviewed 32 participants. The median age of participants was 31; 18 (56%) were MSM and 14 (44%) were FSW. Overall, 12 participants were using PrEP at the time of interview, 9 had previously used PrEP, and 11 had never used PrEP (**Table 1**). Analysis revealed four major themes: (1) social networks as key sources of PrEP information and support; (2) PrEP as protective to individuals and to the community; (3) multiple, but differing, stigmas as barriers to PrEP engagement; and (4) community access as an important strategy to improve PrEP engagement.

**TABLE 1.** Participant Characteristics.

| TABLE 1. Participant Characteristics | KP Category |  |
| --- | --- | --- |
|  | FSW (N=14) | MSM (N=18) |
| **Age Categories** |  |  |
| <25 years | 4 (29%) | 5 (28%) |
| 25-30 years | 2 (14%) | 7 (39%) |
| >30 years | 8 (57%) | 6 (33%) |
| **Education Level** |  |  |
| No school or primary school only | 9 (64%) | 2 (11%) |
| Some secondary school | 4 (29%) | 11 (61%) |
| University | 1 (7%) | 5 (28%) |
| **PrEP use** |  |  |
| Current | 6 (42%) | 6 (33%) |
| Prior | 3 (21%) | 6 (33%) |
| Never | 5 (37%) | 6 (34%) |
| **Sex at birth** |  |  |
| Female | 14 (100%) | 0 (0%) |
| Male | 0 (100%) | 18 (100%) |
| **Sexual Partners** |  |  |
| Men | 14 (100%) | 12 (67%) |
| Men and Women | 0 (0%) | 6 (33%) |
| **Number of Sexual partners in past 6 months** |  |  |
| 1-10 | 1 (7%) | 17 (94%) |
| 11-20 | 4 (29%) | 1 (6%) |
| >20 | 9 (64%) | 0 (0%) |
| **STI diagnosis in prior 6 months** | 7 (50%) | 2 (11%) |
| **Exchange sex for money in prior 6 months** | 14 (100%) | 6 (33%) |
| **Reported challenges related to condom use during sex** |  |  |
| No challenges | 5 (36%) | 9 (50%) |
| Sexual partners refuse condoms because we use PrEP | 0 (0%) | 1 (6%) |
| Condoms feel uncomfortable | 8 (57%) | 3 (17%) |
| Condoms reduce pleasure | 1 (7%) | 5 (28%) |
| **Time since most recent HIV test** |  |  |
| ≤1 month | 1 (7%) | 7 (39%) |
| 2-4 months | 8 (57%) | 8 (44%) |
| ≥5 months | 4 (29%) | 2 (11%) |

### Social networks as key sources of PrEP information and motivation

Overall, participants demonstrated a high level of knowledge about PrEP. Most were aware of its benefit for HIV prevention and understood that it does not protect against other types of STIs, and many described how PrEP could benefit anyone at risk of HIV, in particular KPs. A small number of participants disclosed very limited knowledge about PrEP, not knowing exactly how it was used or confusing it with post-exposure prophylaxis. A number of interviewees reported perceived cost as a barrier to PrEP use, despite it being available for free at health centers in Rwanda. Many participants reported that overall knowledge about PrEP in Rwandan society, and even among the communities of MSM and sex workers, was lacking.

For both MSM and FSW, social networks, including friends, peers, community mobilizers, and KP associations, emerged as key sources of PrEP information, mediated through the trust, acceptance and lack of stigma in these relationships. Participants described a high degree of trust in information that came from friends and peers; individuals who used PrEP felt comfortable sharing information within their communities about its benefits as well as how to access it. The potential for friends and peers to also benefit from PrEP was described as a strong motivation to share information widely, highlighting the key role of these networks as a source of counseling and information. However, many were hesitant to share information about PrEP more broadly because of the potential discrimination they might experience.

> *“I knew it from others because sometimes we talk as friends. So, my friends told me some who are [living with HIV] and those who are [HIV negative]; I used to think that only [people living with HIV] have ART to take but my friends told me that also [people who are HIV-negative] take PrEP due to the fact that some engage in commercial sex which can result into HIV infection. For that reason, I approached a health center for details and the medicine as well, so that I can take it’’*.
>
> *– Female in their 30s, currently on PrEP*
>
> *“I shared it with many people. Most of the time it was to encourage them by saying, ‘It would be great if you take it*.*’…I disclosed it to our community members because it felt normal. I did not share it with non-members of our community*.
>
> *– Male in their 30s, previously on PrEP*

Although most participants felt that the information acquired through social networks was accurate, some described how incorrect (e.g., regarding cost) or negative (e.g., emphasizing side effects) information circulating in these networks discouraged some individuals from engaging in PrEP. One participant described misinformation about uncommon medication side effects:

> *“There exist a lot of rumors that the medicine causes acne and so on. A friend of mine told me that it has caused him facial acne… though I haven’t seen it yet – just rumors – but it’s said that it causes obesity and heart-related effects*.*”*
>
> *– Male in their 30s, currently on PrEP*

Health care providers, including physicians, nurses and community health workers, were felt by most participants to be trustworthy sources of information about PrEP. These sources were particularly important for participants who described difficulty accessing accurate PrEP information through more informal means or who already had an established connection to health centers. As one sex worker explained:

> *We went to there for general treatment, then we heard doctors who were teaching and we were curious to listen to what they were teaching about; we came to find that it was about this PrEP. Then we said, ‘Maybe this is helpful. Let’s proceed*.*’ We approached the nurse, she fully explained it to us, she filed us, and she started giving us the medicine. She helped us*.
>
> *– Female in their 20s, currently on PrEP*

However, for some participants, particularly MSM, stigmatizing attitudes from health care providers overshadowed the perceived and actual benefits of learning about PrEP in health care settings. While participants appreciated the information, counseling and support offered by many health workers, many wished these resources were available in less stigmatizing settings.

> *‘‘Sometimes the health care workers are not aware of particular communities. Let me start with the LGBTI community. You find that [the health care workers] do not have enough information. When they see a man wearing a skirt, they chase the person away without knowing what he seeks and sometimes they do not let him access a service, or they skip him and receive other people due to discriminating against him’’*.
>
> *– Male in their 30s, never on PrEP*

### PrEP protecting both individuals and the community

Among participants currently or previously using PrEP, a major motivator for use was the ease of mind it provided, allowing people to feel more protected from HIV. Participants described how they felt less anxious in terms of the number of partners they had, sexual encounters that were unexpected or with unknown partners, and not needing to negotiate condom use. For sex workers in particular, economic benefits – including taking on additional clients and engaging in higher-paying condomless sex – were seen as substantial incentives for taking PrEP. For example:

> *‘‘The foremost factor is that it’s a protection. The second, it brings inner peace; within this job we tend to worry about the infection but even after unprotected sex you feel safe as long as you’re taking the medicine’’*.
>
> *– Female in their 30s, on PrEP*
>
> *“Most of us sleep with different unfamiliar people and without getting tested. They give us money, and due to life circumstances, you accept. When they ask to have condomless sex, you accept it as well so that your children can eat, but sometimes you take him home and he removes the condom when you did not have an agreement. That is why I take it*.*”*
>
> *– Female in their 20s, previously on PrEP*

A substantial number of participants reflected on the community benefits of PrEP as something that motivated PrEP use. Many described HIV as a serious problem in their communities, one that required an “all hands on deck” approach to address. In this vein, several MSM described how PrEP use would not only benefit themselves, but would benefit sex partners (by indirectly protecting them from HIV), the larger MSM community (by reducing community HIV burden), family members (by reducing the potential shame of being associated with an individual living with HIV), and Rwandan society as a whole. MSM in particular were highly motivated to encourage others in their network to learn more about and use PrEP so that their entire community could benefit. Some examples include:

> *Generally, if someone takes [PrEP], they also protect others from being exposed but when he’s infected, they shall infect others. Therefore, taking the protection also protect others*.
>
> *– Female in their 30s, currently on PrEP*
>
> *If my colleagues took [PrEP] and can prevent themselves [from acquiring HIV], it would be beneficial to me because my colleague can sleep with a seropositive individual and if he doesn’t use the medication, he would contract it and he can transmit it to me if we sleep together… it forms a cycle but if he uses the medication, it would be a solution to my health*.
>
> *– Male in their 30s, never on PrEP*
>
> *“The country itself benefits from PrEP too, because when the number of people living with HIV reduces, the country regains its capacity. As AIDS is a deadly disease, when the mortality cases reduce, a nation retains manpower; hence, human resources will serve their nation. It reduces disputes and trauma in some families, normally when people inform their family that they’re living with HIV, conflicts arise and the family excludes them. Yet, if they are HIV negative…it brings harmony in the family, community and country as a whole*.*”*
>
> *– Male in their 20s, never on PrEP*

### Multiple, but different, stigmas impacting PrEP use for MSM and sex workers

Nearly all participants described HIV-related stigma and fear of discrimination as enormous barriers to PrEP engagement. A dominant theme in interviews was that because there was limited awareness of PrEP in society, and even among some health care providers, PrEP medications were confused with antiretroviral therapy for HIV, and PrEP use would be mistakenly interpreted as having positive HIV status. These fears manifested in various ways: worrying about accidental disclosure of PrEP use if medications were discovered by others; fear of being seen during long waits at a health center, particularly waiting at the HIV program where PrEP is distributed; and anxiety about or direct experience with poor treatment from healthcare providers. Participants also described worry around being perceived as promiscuous if their PrEP status was inadvertently disclosed. Because of these barriers, many individuals took measures to limit disclosure of PrEP use to a limited number of people, mostly sex partners, friends and their medical providers. Two participants described these barriers:

> *‘‘The challenges can be faced by those who use it. The challenges would be to access it rapidly or at a nearby place. Other challenges would be among people who live in families. It can be difficult for them to take it. For them to live with different people, they would discriminate against him, thinking that he is seropositive when he takes a tablet daily*.*”*
>
> *– Male in their 30s, stopped PrEP*
>
> *‘‘Stigmatization is still existing. If someone who look like a girl, goes at a health center and meet a crowd who is waiting for ART or VCT (Voluntary HIV counselling and testing) they stare and gossip about him. Outpatients, other staff such as cleaners have no training about that only care givers do. So, every time the person goes for the service, he’s excluded due to his appearance or identity. Hence saying that stigmatization really exists!”*
>
> *– Male in their 20s, never on PrEP*

For MSM, HIV-related stigma was compounded by stigma around their sexual orientation. MSM participants reported a tremendous amount of anticipated and enacted stigma at health centers, including poor treatment by health care workers. Some described how community health workers living in the same communities as patients, and even medical providers working in health centers, were insufficiently trained on issues of LGBTI health and PrEP, resulting in uncomfortable experiences and explicit discrimination. These mirrored more widespread stigma and discrimination in Rwandan society towards MSM, and made it difficult for some to initiate or continue engaging in PrEP care. As one MSM noted:

> *‘‘So I don’t think that all doctors all over the country have been trained on LGBTI community; even those who were trained, some of them don’t embrace it, consequently they will abuse you. Frankly, if they do not love the community, they’ll not serve you or give you bad service in case of consultation. It discourages us and we can’t dare to come back again, thinking how bad we are treated as if we’re beggars*.*”*
>
> *– Male in their 30s, stopped PrEP*

Participants described other barriers to PrEP use, including competing demands (e.g., work, distance from health center, transportation costs) that made it difficult to attend appointments, side effects leading to discontinuation, or food insecurity that made PrEP a lower priority. For many, not initiating or discontinuing PrEP was not the result of a single factor – rather, the cumulative impact of structural, logistical and stigma-related barriers made the opportunity cost of PrEP use too high. For others, there was sufficient internal or external motivation to overcome stigma, side effects, and other barriers, and continue using PrEP.

### Community access as a critical strategy to improve PrEP engagement

Given the actual and anticipated discomfort and discrimination facing participants, nearly all voiced support for making access to and engagement in PrEP less stigmatizing. Participants in particular expressed their wish to engage in PrEP care in places that were safe and trusted, increase access to information and support, and decrease their exposure to discrimination and related stigma.

Participants advocated for interventions aimed at stigma reduction, such as long-acting PrEP formulations, which would avoid inadvertent disclosure associated with taking daily pills, and changing the packaging of PrEP to clearly differentiate it from antiretrovirals prescribed for HIV. Some participants advocated for PrEP delivery in community settings, which were considered safer, more discreet and easier to access, rather than health centers. These included KP organizations, pharmacies, and even one-on-one delivery by community health workers. Others preferred receiving PrEP care in health centers, which were better resourced and provided more structured care, but felt that additional training of staff on PrEP and LGBTI care was essential to ensuring adequate services.

> “*[Getting PrEP at a community-based, key population association] would reduce the distance covered since it would be close plus in our organization, we understand and know each other, and harassment would not occur, like the challenge I mentioned of finding many people at the health center and as a member of the community you can be afraid to pass there but I think an organization is like home, you cannot be afraid or shy but it would be better if a nurse was available as well to educate us and examine us to know our HIV status before we get the medication’’*.
>
> *– Male in their 20s, currently on PrEP*

## DISCUSSION

In this study of Rwandan MSM and FSW, participants described the key role of social networks as sources of PrEP information and highlighted how peers and friends in these networks helped them overcome PrEP-related barriers and provided motivation to engage in care. Our findings suggest that approaches directed at strengthening key population communities and reducing stigma related to HIV and sexual orientation could facilitate improved, sustained engagement in PrEP.

We found that for both MSM and FSW, community is a central element of PrEP engagement. Key sources of information about PrEP included formal community structures (e.g. associations, peer navigators) as well as less formal friendships and relationships. Participants described a high degree of trust in these sources of information and a corresponding high degree of comfort in accessing them. These findings are similar to research from other SSA settings describing high acceptability of in-person and virtual peer-led communication about PrEP for KPs,^30,31^ indicating that such approaches can be empowering^32^ and even improve engagement in care.^33^ Nonetheless, participants in this study described how erroneous or demotivating information can be amplified by community voices in ways that serve as barriers to PrEP. Countering such misinformation through general and targeted dissemination campaigns – as suggested by participants in this study and others^30,31^ – could help ensure that these trusted sources provide high-quality information.

Participants reported very high levels of stigma and discrimination related to being misidentified as living with HIV and the potential disclosure of their membership in a key population. Both MSM and FSW described HIV-related stigma in their communities (e.g., worry that people would see their medication and think they were using ART) as well as health centers (e.g., being considered promiscuous by health care workers), consistent with findings across SSA.^34,35^ Making access to PrEP easier, more discrete, and less fraught – through long-acting injectable formulations^36^ and low-barrier, community-based access points – was preferred by many participants. For MSM, HIV-related stigma was compounded by stigma related to their sexual orientation, which was felt to be pervasive in society including in many health centers, consistent with national survey data from Rwanda.^37^ Because of this, many expressed a desire for PrEP delivery in LGBTI community organizations, similar to preferences identified by MSM in other SSA settings.^31^ Notably, not all participants felt that community-based PrEP delivery options were ideal, with some preferring more formal and better resourced health settings. Recent data from the SEARCH trial in Uganda and Kenya indicate that providing individuals with choices (e.g., service location) can increase PrEP coverage.^38^ While that study did not report on sexual orientation and likely included few MSM or FSW, our data suggest that offering options for PrEP care would be well received by key populations in SSA.

Most studies of KPs in SSA to date have examined the individual benefits of, facilitators of and barriers to PrEP, yet few have assessed the community aspects of prevention. We observed a high degree of community-focused PrEP motivation among both MSM and FSW participants, who identified HIV as a serious problem in their social networks and described how PrEP engagement could benefit sex partners, family members, and peers. These findings echo the concepts of collective antiretroviral protection and prevention solidarity, recently described by Brisson, et al.^39^ Much like the idea of herd immunity – where unvaccinated individuals in a population benefit from the vaccination of others – collective protection suggests that when a substantial portion of a community remains HIV-negative (or, if living with HIV, virally suppressed), all members of the community have a reduced HIV risk. Although participants in this study did not explicitly describe the concept of collective antiretroviral protection, our results suggest that some MSM and FSW in Rwanda are actively considering how their individual PrEP-related decisions can benefit others around them. Rwandan society has been considered one with a high degree of social cohesion,^40^ and it is possible that prevention solidarity might have less potential in other settings. Nonetheless, collective HIV prevention has been described among heterosexual, serodiscordant couples in Uganda and Kenya, although interpersonal dynamics may differ substantially in these relationships compared to those in broader social networks of MSM and FSW.^41,42^ To our knowledge, prevention solidarity has not been examined among MSM or FSW in SSA. Understanding how to best implement collective antiretroviral protection among MSM and FSW in SSA could be a key step to improving and maintaining PrEP engagement.

While analysis revealed many common themes among MSM and FSW, we found important differences as well. For MSM, anticipated and experienced sexual stigma in society as well as in health centers led to the social network emerging as the single most important source of information, motivation and support related to PrEP engagement. These findings are consistent with prior research we conducted highlighting the close-knit nature of these networks,^22^ and support recent PEPFAR recommendations to strengthen LGBTI associations to ensure dissemination of accurate PrEP information and facilitate peer linkage to PrEP.^27^ FSW, in contrast, described more informal social networks and were more comfortable accessing care in health centers. Many FSW also emphasized economic motivations for PrEP use and highlighted logistical barriers. Together, these findings suggest that efforts to disseminate PrEP information and motivate engagement among MSM and FSW should not follow a uniform approach, and to the degree possible should be tailored to the needs of each community, aligned with calls for differentiated approaches to HIV prevention.^43^

This study has several limitations. We interviewed a sample of MSM and FSW in a single city in SSA with a relatively tolerant policy climate with respect to sexual health. Therefore, our findings may not be representative of key populations who live in rural areas and in other SSA countries. Some interviews occurred in health care settings, and it is possible that participants may have felt reluctant to fully describe their perspectives on health care because of social desirability bias. Willingness to participate in research may reflect an overall lower degree of stigma, and we therefore may not have captured the perspectives of KP who are too stigmatized to enroll in a research study.

In conclusion, we found that for MSM and FSW in Rwanda, despite high levels of PrEP awareness in their communities, PrEP engagement remains challenging because of the potential to be misidentified as having HIV and because of stigma related to membership in a key population group. Nonetheless, participants highlighted the strength and support they experienced from peers and colleagues in their social networks, and described a strong motivation to use PrEP as a way to protect both themselves and their communities from HIV. Our findings suggest that leveraging community resources for disseminating information about HIV prevention and delivering PrEP could contribute to successful implementation of PrEP for MSM and FSW in Rwanda and other settings in SSA.

## Data Availability

All data produced in the present study are available upon reasonable request to the authors

## DECLARATIONS

### Ethical approval and informed consent

All research participants provided written informed consent prior to initiating research procedures. Ethical approval was obtained from the Rwanda National Ethics Committee and the Institutional Review Board of the Albert Einstein College of Medicine.

### Consent for publication

Not applicable

### Availability of data and materials

The datasets analyzed during the current study are available from the corresponding author on reasonable request.

### Authors contributions

JR, AA and VP conceived of and led the study and oversaw data analysis; GM contributed to study design. JG, GM and FM were responsible for data collection and cleaning. JR and JG led data analysis, with support from GM, FM and NZ. JR, JG, NZ, AA, GM and VP contributed to interpretation of study data. All authors read and approved the final manuscript.

## Acknowledgments

We are especially grateful for the participants who provided their time and insight.

